# Social Isolation among Mothers Caring for Infants in Japan: Findings from the Nationwide Survey of Healthy Parents and Children 21

**DOI:** 10.1101/2020.11.12.20230839

**Authors:** Sayaka Yamazaki, Yuka Akiyama, Ryoji Shinohara, Zentaro Yamagata

## Abstract

**Background:** Child-rearing isolation may increase the risk of child abuse and negatively affect child development owing to increased urbanization and decline in family and community support systems.

**Purpose:** This study aimed to identify the prevalence of child-rearing isolation and the related sociodemographic factors among mothers in Japan using data from the Final Survey of Healthy Parents and Children 21.

**Participants:** Mothers of young children attending their health checkups.

**Methods:** Multivariate logistic regression models assessed the association between child-rearing isolation and socio-demographic variables. Data from 69,337 women were analyzed.

**Results:** Mothers who experienced child-rearing isolation comprised 0.2% of all participants. Mothers who were 35- to 39-years-old at childbirth (OR = 1.6, CI[1.0, 2.4], *p* = .036), unemployed (OR = 1.7, CI[1.3, 2.4], *p* = .001), experienced financial difficulty (OR = 1.8, CI[1.3, 2.5], *p* < .001), had husbands with limited participation in child-rearing (OR = 5.7, CI[4.2, 7.9], *p* < .001), lived in special wards in Tokyo Metropolis (OR = 4.2, CI[2.2, 8.3], *p* < .001), had child abuse concerns (OR = 2.1, CI [1.5, 2.9], *p* < .001), and had no time to relax with their child (OR = 4.5, CI [3.1-6.7], *p* < .001) exhibited higher odds ratio for child-rearing isolation, compared to those who did not.

**Conclusions:** Findings showed the impact of urban living on maternal health, the influence of isolation on mothers’ anxiety about child-rearing and their potential for child maltreatment. The importance of fathers’ involvement in child-rearing for preventing maternal child-rearing isolation was highlighted.

**Significance:** Previous studies have shown that social isolation is significantly associated with morbidity and mortality. One of the most pressing issues in Japan is child-rearing isolation of mothers with infants. Child-rearing isolation is considered a risk factor for abuse and disruption of healthy parent-child relationships. However, no studies have examined the associated factors of child-rearing isolation among mothers, adjusting for confounding factors. Our results provide evidence that child-rearing isolation is a risk factor for maltreatment. In addition, we found new evidence that maternal child-rearing isolation is significantly associated with age at birth, lack of spousal participation in parenting, and urban living.

## Introduction

Social isolation has been identified as a risk factor for morbidity and mortality for a number of diseases (Elovainio et al., 2017; Leigh-Hunt et al., 2017). Therefore, preventing social isolation is one of the key challenges that public health faces.

Social isolation can occur in any age group; however, *child-rearing isolation* of mothers with infants is a pressing issue in Japan. In fact, it is addressed as a major issue in three maternal and child health policies in Japan: countermeasures against adverse effects of the relationship between parents and children (Maternal and Child Health Division, Children and Families Bureau, Ministry of Health, Labour and Welfare, 2000), child abuse (Ministry of Health, Labour and Welfare, 2005), and birth rate decline (Cabinet Office, 2004). In previous studies, social isolation has been defined as the subjective feeling about one’s life lacking social connections or social support (Holt-Lunstad et al., 2015), as well as a situation in which one lacks anyone to consult regarding important matters (Brashears, 2011). Specifically, maternal social isolation has been defined as mothers lacking anyone to speak or consult with regarding child-rearing concerns when they struggle with child-rearing (Honda et al., 2019).

The lack of social support has been shown to be associated with maternal stress and depression (Mulvaney and Kendrick, 2005a) and not adopting safe practices for the prevention of childhood injury (Mulvaney and Kendrick, 2005b). Moreover, child-rearing isolation represents a risk factor for child abuse (U.S. Department of Health and Human Services, 2019), which has a deleterious impact on children’s development. In Japan, socially isolated mothers with 6-month-old infants spent JPY 4,186 (Japanese yen; around USD 35) more per month on child-rearing costs compared to non-isolated mothers (Honda et al., 2019), which suggested parenting costs are higher due to lack of social support for mothers. Therefore, from a public health perspective, understanding the current situation regarding child-rearing isolation and its related factors is critical. However, to date, few studies in Japan have examined child-rearing isolation nationwide.

Therefore, this study aims to examine the relationship between child-rearing isolation and socio-demographic variables including maternal age, child age, employment, socioeconomic status, spousal support, living municipality, and maternal psychosocial wellbeing using data from all prefectures in the Final Survey of Healthy Parents and Children 21 (Yamagata et al., 2014).

## Methods

### Setting and Participants

Healthy Parents and Children 21 is a national campaign to promote maternal and child health in Japan (Osawa et al., 2019), which conducted a final survey to reveal new issues in 2014. This study used the data from the final survey as a secondary analysis of an existing data set.

Participants were residents of 472 municipalities that had been identified as targets of the Final Survey of Healthy Parents and Children 21 and were also the parents of children scheduled for child health checkups during the survey period. Municipalities were divided into quartiles by population, and participants were randomly selected from each group. In total, 89,404 print questionnaires were mailed to the identified number of participants in each municipality. Surveys were collected from 75,622 parents whose children were receiving health checkups during the survey period: 3-4-month checkup, *n* = 20,729; 18-month checkup, *n* = 27,922; 3-year checkup, *n* = 26,971 (response rate = 84.6%). The surveys were mailed in February 2013 and were collected between April and August 2013. Surveys with missing variables were excluded. Therefore, the final sample size for this study was 69,337 mothers (Figure 1).

**Figure 1.**
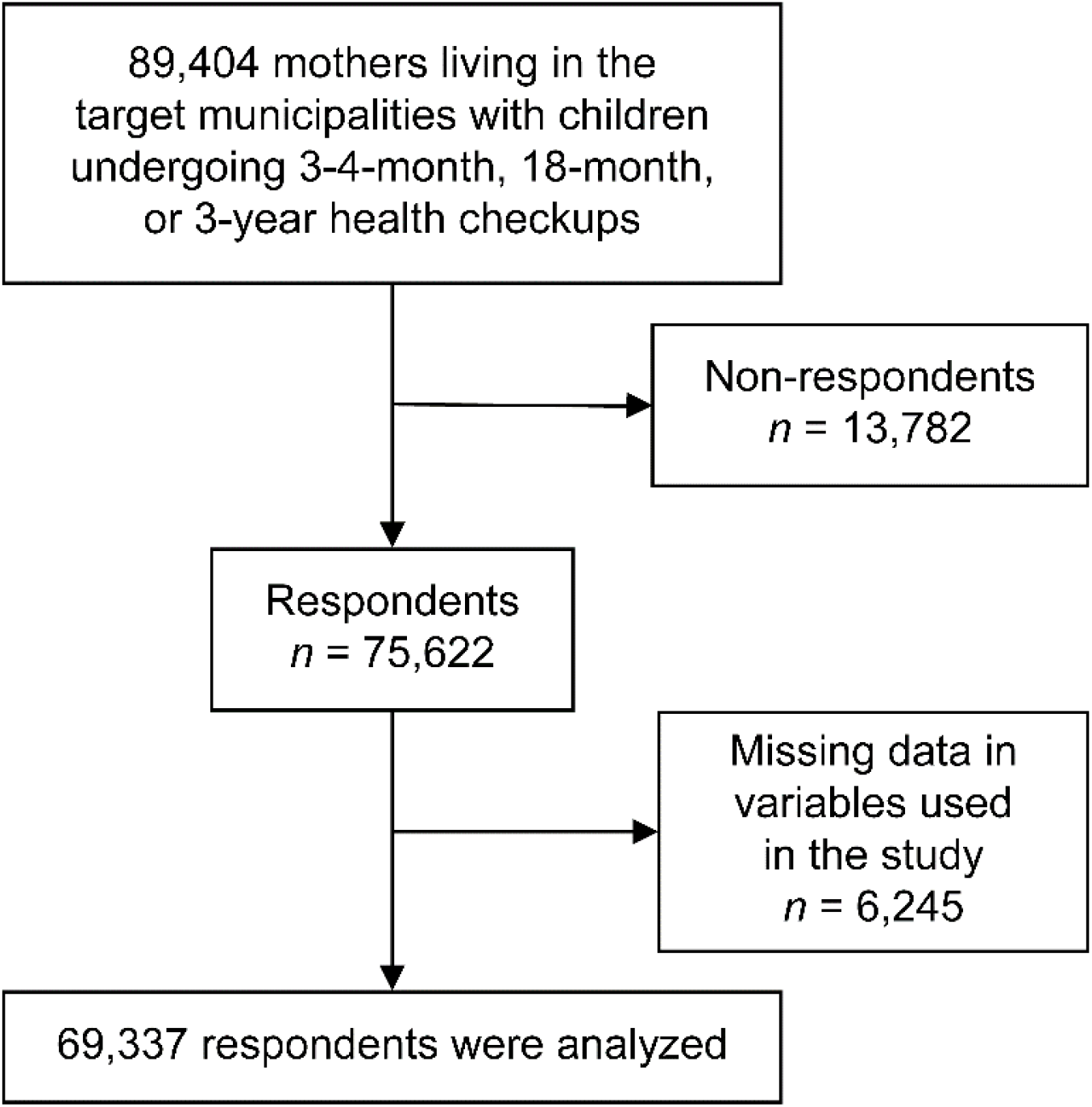
Flowchart of study participants.

### Sampling and Data Collection

The Maternal and Child Health Section of each municipality requested parents of young children who were scheduled for health checkups to complete the “Parents and Children Health Survey Questionnaire,” which was collected at the children’s checkups. This questionnaire was created to clarify how much the mental and physical health of children and parents had improved for the identified issues surrounding maternal and child health in Healthy Parents and Children 21.

### Included Variables

The dependent variable in this study was maternal child-rearing isolation. This was measured by the question, “who is the person you [the mother] consult about your child-rearing in daily life?” and mothers were asked to choose from 11 options (multiple answers allowed), including: *my husband, the child’s grandparents, neighbor(s), friend(s), primary doctor, a public health nurse or midwife, nursery or preschool teachers, telephone consultation, the Internet, other*, and *nobody*. We identified the mothers who were “isolated in child-rearing” as those who responded *nobody* to the aforementioned question. Mothers who selected any of the other ten options were defined as being “not-isolated in child-rearing.” The use of the Internet, especially social networking sites, is associated with having social networks outside of one’s family and it is related to the formation of weak ties at the neighborhood level (Hampton et al., 2011); therefore, we considered the Internet as a potential consultant in this study.

The explanatory variables were selected based on existing studies regarding factors associated with child-rearing isolation in mothers with infants (Honda et al., 2019). To further explore the demographic and psychological factors associated with maternal isolation, we added the “anxiety about child-rearing” and “living municipality” items. The selected variables included the demographic characteristics of the mother, the age of the child, the environmental factors associated with child-rearing, and child-rearing anxiety. The selected explanatory variables included the maternal age at childbirth (< 25, 25–29, 30–34, 35–39, and > 39 years), child’s age (3–4 months, 18 months, and 3 years), mother’s employment status (employed or unemployed), socioeconomic status (average or above average vs. difficult), husband’s participation in child-rearing (yes vs. almost none), living municipality (city, city designated by government, town/village, and special ward: Tokyo ward), time to relax with the child (yes vs. no), child-rearing confidence (confident vs. not confident), and concerns regarding abusing the child (yes vs. no).

### Statistical Analysis

A multivariate logistic regression analysis by means of a forced entry method was performed using complete data with no missing variables. Further, for sensitivity analysis, we accounted for missing data with the multiple imputation by chained equations (MICE) for two hundred imputed datasets (White et al., 2011). The imputation model included all the study variables. In each case, the odds ratio and 95% confidence intervals were calculated, and the statistical significance level was set at *p* < .05. Stata ver. 13 (StataCorp LP, College Station, Texas, USA) was used as the software program for all analyses.

### Ethical Considerations

As an ethical consideration, the questionnaires were completed anonymously without any form of identifying information. As this is a secondary analysis of an existing data set with no access to personal identifiers, the requirement for informed consent was waived. Data analysis and publication of the results were approved by the Ethics Committee at XXX (receipt number: XXX).

## Results

### Demographic Characteristics

Table 1 shows the participants’ characteristics. Of the 69,337 mothers who provided complete data, 160 (0.2%) reported child-rearing isolation. Among the mothers reporting child-rearing isolation, 26.9% (*n* = 43) were 35 to 39-years-old, compared to only 20.4% (*n* = 14,094) of the non-isolated mothers being in that age group. Moreover, 5.6% (*n* = 9) of the isolated mothers were 40 years old, compared to 3.3% (*n* = 2,264) of the non-isolated mothers.

**Table 1.**
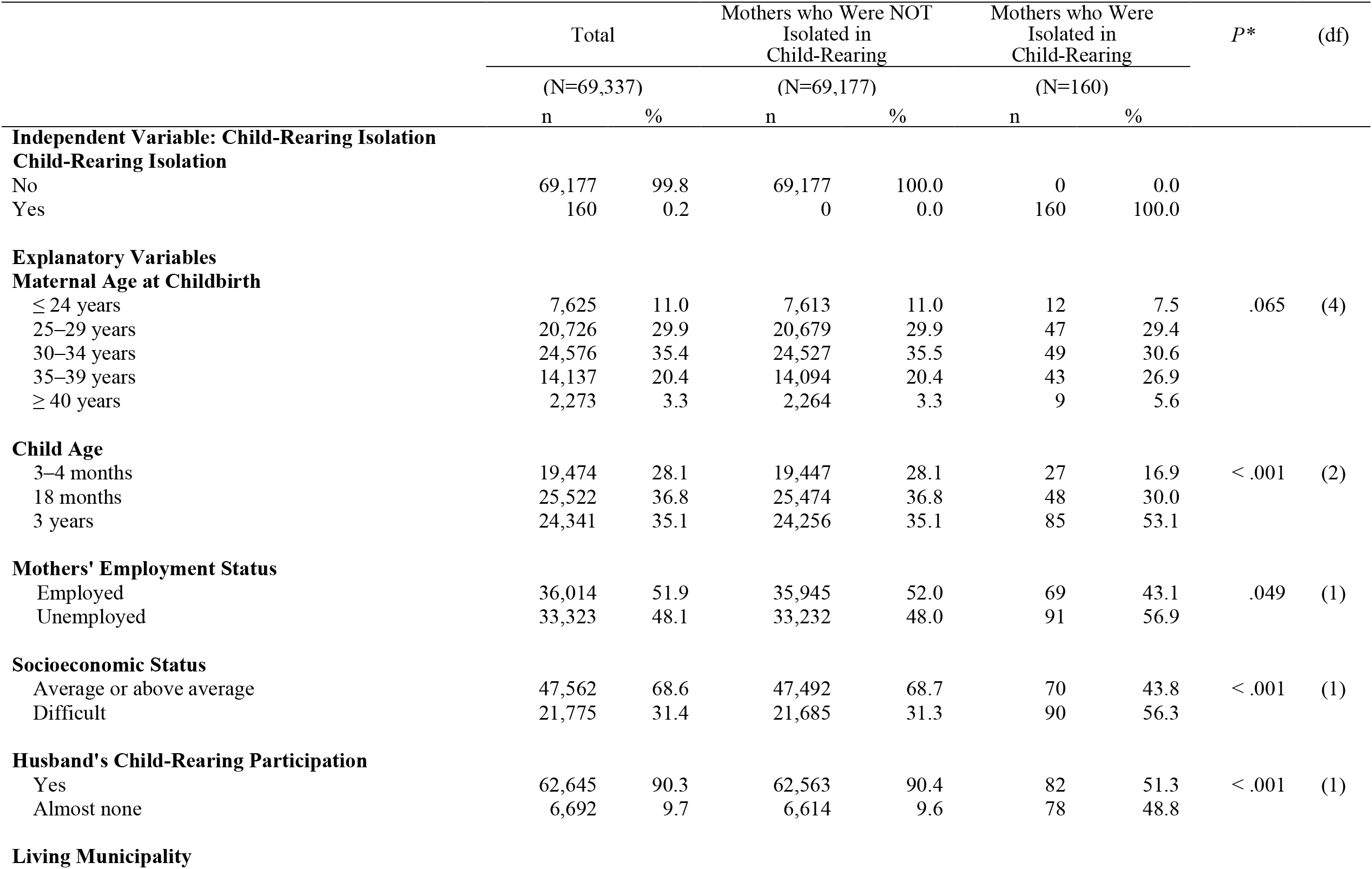
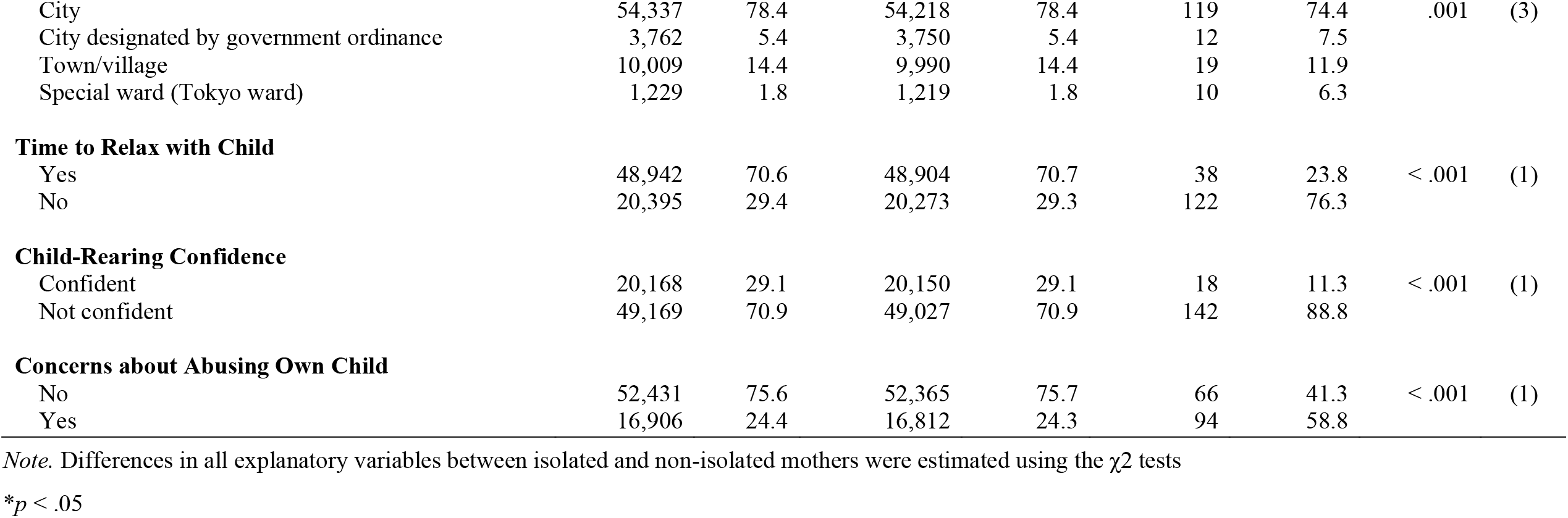
Participants’ Characteristics.

The majority of mothers who reported child-rearing isolation had 3-year-old toddlers (*n* = 85, 53.1%), while this was reported by only 35.1% of the non-isolated mothers (*n* = 69,177). Moreover, more than half of the isolated mothers were unemployed (*n* = 91, 56.9%), whereas in non-isolated mothers the results were more evenly distributed, with slightly more mothers being employed (*n* = 35,945, 52.0%) than unemployed (*n* = 33,232, 48.0%). This finding demonstrates a reverse trend from the overall sample, as 48.1% (*n* = 33, 323) of all mothers were unemployed while 56.9% (*n* = 91) of the isolated mothers were unemployed.

The overwhelming majority (*n* = 141, 88.2%) of isolated mothers lived in an urban environment (i.e., city, city designated by government ordinance, or special ward); which is analogous to their proportion of the total sample size, as they represented 85.6% (*n* = 59,187) of the non-isolated mothers. Those living in the special wards in Tokyo were over-represented among the socially isolated mothers, as they represented 1.8% (*n* = 1,219) of the non-isolated mothers but 6.3% (*n* = 10) of the isolated mothers.

Although 90.4% (*n* = 62,563) of the non-isolated mothers reported that the children’s fathers helped with child-rearing, only approximately half of the isolated mothers reported receiving assistance from the fathers (*n* = 82, 51.3%). In addition, 76.3% (*n* = 122) of the isolated mothers reported not having time to relax with their children; compared to only 29.3% (*n* = 20,273) of the non-isolated mothers. Further, 88.8% (*n* = 142) of the socially isolated mothers reported lacking confidence about their child-rearing, compared to only 24.3% (*n* = 16,812) of the non-isolated mothers. Moreover, 58.8% (*n* = 94) of the socially isolated mothers reported being concerned about abusing their children.

Mothers who reported experiencing financial difficulties comprised only 31.3% (*n* = 21,685) of the non-isolated mothers but 56.9% (*n* = 90) of the isolated women; thus, mothers with financial concerns were overrepresented in the socially isolated group.

### Logistic Regression Analysis Results

A multiple logistic regression analysis was conducted. A significant overall regression equation was found, χ^2^(15) = 354.84, *p* < .001, with a McFadden pseudo *R*^2^ = .16. Table 2 shows the results of the logistic regression analysis (univariate/multivariate).

**Table 2.** Results of Logistic Regression Analysis with “Isolation of Child-Rearing” as the Objective Variable.

|  | Univariate analysis |  |  |  | Multivariate analysis |  |  |
| --- | --- | --- | --- | --- | --- | --- | --- |
|  | n | OR | 95% CI | (N=69,337)<br>*P* * | OR | 95% CI | *P* * |
| **Maternal Age at Childbirth** |  |  |  |  |  |  |  |
| ≤ 24 years | 7,625 | 0.8 | [0.4, 1.5] | .462 | 0.8 | [0.4, 1.5] | .479 |
| 25–29 years | 20,726 | 1.1 | [0.8, 1.7] | .528 | 1.2 | [0.8 ,1.8] | .331 |
| 30–34 years | 24,576 | ref |  |  |  |  |  |
| 35–39 years | 14,137 | 1.5 | [1.0, 2.3] | .043 | 1.6 | [1.0, 2.4] | .036 |
| ≥ 40 years | 2,273 | 2.0 | [1.0, 4.1] | .058 | 1.9 | [0.9, 3.9] | .078 |
| **Child Age** |  |  |  |  |  |  |  |
| 3–4 months | 19,474 | ref |  |  |  |  |  |
| 18 months | 25,522 | 1.4 | [0.8, 2.2] | .205 | 1.0 | [0.6, 1.6] | .979 |
| 3 years | 24,341 | 2.5 | [1.6, 3.9] | < .001 | 1.5 | [1.0, 2.4] | .071 |
| **Mothers' Employment Status** |  |  |  |  |  |  |  |
| Employed | 36,014 | ref |  |  |  |  |  |
| Unemployed | 33,323 | 1.4 | [1.0, 2.0] | .026 | 1.7 | [1.3, 2.4] | .001 |
| **Socio-Economic Status** |  |  |  |  |  |  |  |
| Average or above average | 47,562 | ref |  |  |  |  |  |
| Difficult | 21,775 | 2.8 | [2.1, 3.8] | < .001 | 1.8 | [1.3, 2.5] | < .001 |
| **Husband's Participation in Child-Rearing** |  |  |  |  |  |  |  |
| Yes | 62,645 | ref |  |  |  |  |  |
| Almost none | 6,692 | 9.0 | [6.6, 12.3] | < .001 | 5.7 | [4.2, 7.9] | < .001 |
| **Living Municipality** |  |  |  |  |  |  |  |
| City | 54,337 | ref |  |  |  |  |  |
| City designated by government ordinance | 3,762 | 1.5 | [0.8, 2.6] | .214 | 1.3 | [0.7, 2.4] | .378 |
| Town/village | 10,009 | 0.9 | [0.5, 1.4] | .562 | 0.8 | [0.5, 1.4] | .491 |
| Special ward (Tokyo ward) | 1,229 | 3.7 | [2.0, 7.1] | < .001 | 4.2 | [2.2, 8.3] | < .001 |
| **Time to Relax with Child** |  |  |  |  |  |  |  |
| Yes | 48,942 | ref |  |  |  |  |  |
| No | 20,395 | 7.7 | [5.4, 11.2] | < .001 | 4.5 | [3.1, 6.7] | < .001 |
| **Child-Rearing Confidence** |  |  |  |  |  |  |  |
| Confident | 20,168 | ref |  |  |  |  |  |
| Not confident | 49,169 | 3.2 | [2.0, 5.3] | < .001 | 1.4 | [0.9, 2.4,] | .174 |
| **Concerns about Abusing Own Child** |  |  |  |  |  |  |  |
| No | 52,431 | ref |  |  |  |  |  |
| Yes | 16,906 | 4.4 | [3.2, 6.1] | < .001 | 2.1 | [1.5, 2.9] | < .001 |

\* $p < .05$

The results of the multivariate logistic regression analysis showed that the mothers whose age at childbirth was 35- to 39-years-old had significantly higher odds of reporting child-rearing isolation compared to those whose ages at childbirth were 30- to 34-years-old (OR = 1.6, 95% CI[1.0, 2.4], *p* = .036). Mothers who were unemployed (OR = 1.7, 95% CI[1.3, 2.4], *p* = .001), reporting financial difficulties (OR = 1.8, 95% CI[1.3, 2.5], *p* < .001), and whose husbands did not offer significant support in the child-rearing (OR = 5.7, 95% CI[4.2, 7.9], *p* < .001), showed significantly higher odds for having child-rearing isolation compared to those in the reference groups (Table 2). Regarding the municipalities, mothers living in special wards (i.e., the wards of Tokyo Metropolis) had significantly higher odds of reporting child-rearing isolation than those living in other cities (OR = 4.2, 95% CI[2.2, 8.3], *p* < .001). Mothers who reported having concerns regarding abusing their children (OR = 2.1, 95% CI[1.5, 2.9], *p* < .001) and having no time to relax with their children (OR = 4.5, 95% CI[3.1-6.7], *p* < .001) had higher odds of reporting social isolation compared to those mothers who were not concerned about abusing their children and reported having time to relax with them. There were statistically significant differences in most variables between those who were and those who were not included in this analysis. Details are presented in Appendix 1. However, in the sensitivity analysis, there were small differences between the results of the analysis of the complete data and those of the MICE. Details are presented in Appendix 2. Frequencies of complete and imputed variables are reported in Appendix 3 (imputed data sample size 75,622).

## Discussion

We identified a number of variables that were significantly associated with child-rearing isolation based on the results of the multivariate logistic regression analysis (Table 2).

### Association with Maternal Age at Childbirth

Mothers whose age at childbirth was 35- to 39-years-old had significantly higher odds for child-rearing isolation compared to those aged 30–34 at childbirth, suggesting that being older at the time of childbirth is associated with child-rearing isolation. Previous research examined adolescent mothers’ isolation (Kim et al., 2017), however, few studies have investigated the association between advanced maternal age and the child-rearing environment. According to a report by the Organisation for Economic Cooperation and Development (OECD; OECD Family Database, 2019), most women giving birth to their first child are age 30 or older in OECD counties, and the average age of women at childbirth has increased by 2 to 5 years from 1970 to 2017. Since advanced maternal age at childbirth will likely increase in the future, risk factors, including the child-rearing environment as well as pregnancy outcomes, associated with it should be explored in future research.

### Association with Maternal Employment

Compared to working mothers, non-working mothers had significantly higher odds of reporting child-rearing isolation. In a previous study, the absence of opportunities for social interaction in the workplace was listed as one of the major structural causes of social isolation (Stewart et al., 2009), and researchers have identified paid jobs as a critical area of social contact and interaction (Gordon et al., 2000). Thus, unemployed mothers are presumed to have relatively fewer opportunities for social interaction and fewer social support resources compared to employed mothers.

Non-working mothers spend a greater percentage of their time with their children and had relatively fewer social interactions with others compared to working mothers, which might lead to child-rearing isolation. Although maternal employment serves as a factor to prevent child-rearing isolation, research has suggested that mothers must involuntarily quit their jobs in some situations to assume their roles as mothers (Mitsubishi UFJ Research and Consulting, 2015); thus, improvements in the working environment that promote mothers with infants returning to work while raising their children are desirable.

### Socio-Economic Status

Mothers who reported having financial difficulties had significantly higher odds of child-rearing isolation compared to those who reported having average or above average finances. Previous studies have shown that those with a low-income have an increased risk of social isolation (Gallie et al., 2003). Since economic disadvantages such as poverty and unemployment are associated with shame and stigma, individuals with these challenges might restrict or avoid social contact (Lindsay, 2010).

Stewart et al. (2009) reported that individuals with low household incomes had extremely limited social support resources compared to those reporting high incomes. Structural factors contributing to social isolation for those living with a low-income include lacking the financial means to access public facilities, lacking social interaction due to insufficient education and employment opportunities, avoiding families and colleagues due to poverty, and engaging in self-isolation when fearing the threat of criticism (Stewart et al., 2009). Isolated mothers spend more money on child-rearing than non-isolated mothers (Honda et al., 2019), which may lead to financial difficulty. Overall, these results may suggest that financial difficulties might result in child-rearing isolation, and child-rearing isolation leads to increased child-rearing cost, which creates a vicious cycle.

### Association with Husband’s Child-Rearing

In a study conducted in the United States, individuals that reported having a network of people who are not relatives that they can talk with regarding important matters were one-third smaller than those reported in 1985 (McPherson et al., 2006). In Japan, mothers consult primarily with their husbands about child-rearing on a daily basis (i.e., emotional support source; Yamazaki et al., 2018). Thus, given the current trends, the social ties with those who are not relatives have decreased, and intimate networks are primarily comprised of spouses. A similar change has been observed among mothers with infants, suggesting that when no spousal social support is available, mothers can easily become isolated. Therefore, lacking social support from husbands is a factor directly linked to child-rearing isolation in mothers.

UNICEF has indicated that although Japan’s parental leave system is substantial, the percentage of fathers taking paternity leave is low (UNICEF, 2019a). A report by the International Network on Leave Policies and Research (2019), reported that percentage of fathers taking up paternal leave between 2015 and 2018 varied across countries. The reasons for not taking paternal leave for Japanese men include a limited workforce and an unsupportive atmosphere in the workplace, which accounted for a high percentage of men being hesitant to take time off (Cabinet Office, 2018). Much evidence has demonstrated that paternity leave increases fathers’ involvement in child-rearing, which is beneficial for both infants and mothers (UNICEF, 2019b), suggesting that paternity leave should be provided and supported by employers.

### Associations with Place of Residence

Mothers living in Tokyo special wards had significantly higher odds of child-rearing isolation compared to those living in other places. Due to the unipolar concentration of Japan’s politics and economy, in Tokyo’s special wards, newly developed residential areas and high-rise apartments have increased. It has been reported that mothers with infants consider establishing friendships and social support networks a “time-consuming process” in newer residential areas (Strange et al., 2016). Mothers living in a metropolis are more likely to increase their risk for child-rearing isolation until a support network is built in the metropolis.

The proportion of the world’s population living in urban areas is increasing. Currently, half of the world’s population lives in urban cities and it is expected to increase to 70% by 2050 (Kennedy and Adolphs, 2011). The urban environment has been found to increase the risk of mental disorders (Krabbendam and Van, 2005; Lederbogen et al., 2011; Pedersen and Mortensen, 2001), as well as the risks for hypertension, overweight, and diabetes (Eckert and Kohler, 2014). Although urban life might increase various health-related risks, little data has linked urban features such as isolation to population health. It is necessary to consider policies taking into account the high risk of child-rearing isolation for mothers living in an urban environment.

### Association with Child-Rearing Anxiety

Variables related to anxiety about child-rearing was significantly associated with child-rearing isolation. Child-rearing isolation represents a risk factor for child abuse (U.S. Department of Health and Human Services, 2019), and the parenting environment of having no one to talk to about the child has been reported to be significantly associated with child abuse (Mochizuki et al., 2014). Our research complements findings from previous studies, which reported an association between child-rearing isolation and child abuse.

Increased anxiety has been shown to lower the desire for social contact (Sarnoff and Zimbardo, 1961). Thus, child-rearing anxiety, such as concerns regarding abusing the child, might lead mothers to avoid social contact. In that sense, a mother who has concerns about her own parenting may be isolated without anyone to talk to, which may result in child abuse. Additionally, social isolation has been reported to be associated with increased mental health risks, including mood disorders (Chou et al., 2011). These mental health issues might mediate the association between child-rearing isolation and anxiety. Early detection and the prevention of child-rearing isolation might alleviate mother’s anxiety about child-rearing and prevent child abuse and mental health challenges. Therefore, developing child-rearing support systems that can prevent isolation of child-rearing mothers should be a priority.

### Limitations and Possibilities of this Study

There has not been a unified definition of social isolation in previous studies, therefore, our results are dependent on the definition used in this study. Missing data reduced the sample size at a multivariate logistic regression analysis, although the results were similar when using missing data imputed with MICE. The possibility of residual confounding cannot be completely ruled out in observational studies such as ours. Since this research followed a cross-sectional design, drawing causal inferences is not possible, as the directionality between child-rearing isolation and the associated variables may be reversed.

This study’s main strength is that it clarified the actual situation of mothers’ child-rearing isolation in Japan and examined the associated factors using large, nationally representative data. A large sample size substantially reduced the risk of random error and allowed us to have adequate power for analysis. In addition, while the child-rearing isolation percentage is low, child-rearing isolation still has detrimental effects on child development and maternal health, making this study extremely significant from an epidemiological standpoint.

### Future Perspectives

Mothers who are isolated in child-rearing may have many risk factors, such as poverty, child-rearing anxiety, lack of support from the spouse, and, particularly, child abuse; thus, identifying sources of support to eliminate child-rearing isolation is necessary. Therefore, the degree of father’s involvement in child-rearing is thought to contribute largely to child-rearing isolation in mothers. Improvements in the work environment to allow both mothers and fathers to continue to work while raising their children are needed. Public health policies addressing these issues might reduce adverse health effect for mothers, children, and fathers. Therefore, to clarify the association between isolation and health outcomes, longitudinal and intervention studies are needed in the future.

## Conclusions

We examined the characteristics of mothers reporting child-rearing isolation and the associated factors using data from a large-scale, nationwide survey in Japan. Child-rearing isolation was found to be associated with advanced age at childbirth, unemployment, financial difficulties, lack of husband’s participation in child-rearing, anxiety about child-rearing: concerns about abusing own child, no time to relax with child, and living in Tokyo’s special wards.

## Data Availability

The data that support the findings of this study are available from the corresponding author, [Sayaka Yamazaki], upon reasonable request.

## Acknowledgements

This study was supported by Health Science and Labor Research Grants, Japan, Project for Baby and Infant in Research of Health and Development to Adolescent and Young Adult, “Study on Utility of Maternal and Child Health Information for Improvement of Maternal and Child Health” (Principal Investigator: Zentaro Yamagata; Project number: H-28-Sukoyaka-Ippan-001, 19DA1003). The current study was conducted using the data collected in the study funded by that grant. Independent of the study funded, the authors designed this study, analyzed data, interpreted it, wrote the article and decided to submit it for publication.

## Ethical Considerations

As an ethical consideration, the questionnaires were completed anonymously without any form of identifiable information. As this is a secondary analysis of an existing data set with no access to personal identifiers, the requirement for informed consent was waived. Data analysis and publication of the results were approved by the Ethics Committee at Yamanashi University Faculty of Medicine (receipt number: 1119).

**Appendix 1.** Differences Between Included and Excluded Participants.

|  | Included |  | Excluded |  | *P* * | (df) |
| --- | --- | --- | --- | --- | --- | --- |
|  | *n* | % | *n* | % |  |  |
| **Independent Variable: Child-Rearing Isolation** |  |  |  |  |  |  |
| **Child-Rearing Isolation** |  |  |  |  |  |  |
| No | 69,177 | 99.8 | 3,416 | 99.4 | < .001 | (1) |
| Yes | 160 | 0.2 | 19 | 0.6 |  |  |
| **Explanatory Variables** |  |  |  |  |  |  |
| **Maternal Age at Childbirth** |  |  |  |  |  |  |
| ≤ 24 years | 7,625 | 11.0 | 1,272 | 21.4 | < .001 | (4) |
| 25–29 years | 20,726 | 29.9 | 1,678 | 28.2 |  |  |
| 30–34 years | 24,576 | 35.4 | 1,747 | 29.4 |  |  |
| 35–39 years | 14,137 | 20.4 | 1,000 | 16.8 |  |  |
| ≥ 40 years | 2,273 | 3.3 | 243 | 4.1 |  |  |
| **Child Age** |  |  |  |  |  |  |
| 3–4 months | 19,474 | 28.1 | 1,255 | 20.0 | < .001 | (2) |
| 18 months | 25,522 | 36.8 | 2,400 | 38.2 |  |  |
| 3 years | 24,341 | 35.1 | 2,630 | 41.8 |  |  |
| **Mothers' Employment Status** |  |  |  |  |  |  |
| Employed | 36,014 | 51.9 | 2,398 | 64.2 | < .001 | (1) |
| Unemployed | 33,323 | 48.1 | 1,335 | 35.8 |  |  |
| **Socioeconomic Status** |  |  |  |  |  |  |
| Average or above average | 47,562 | 68.6 | 2,072 | 56.0 | < .001 | (1) |
| Difficult | 21,775 | 31.4 | 1,628 | 44.0 |  |  |
| **Husband's Child-Rearing Participation** |  |  |  |  |  |  |
| Yes | 62,645 | 90.3 | 1,493 | 88.8 | .030 | (1) |
| Almost none | 6,692 | 9.7 | 189 | 11.2 |  |  |
| **Living Municipality** |  |  |  |  |  |  |
| City | 54,337 | 78.4 | 5,090 | 81.0 | < .001 | (3) |
| City designated by government ordinance | 3,762 | 5.4 | 242 | 3.9 |  |  |
| Town/village | 10,009 | 14.4 | 882 | 14.0 |  |  |
| Special ward (Tokyo ward) | 1,229 | 1.8 | 71 | 1.1 |  |  |
| **Time to Relax with Child** |  |  |  |  |  |  |
| Yes | 48,942 | 70.5 | 2,453 | 66.6 | < .001 | (1) |
| No | 20,395 | 29.2 | 1,228 | 33.4 |  |  |
| **Child-Rearing Confidence** |  |  |  |  |  |  |
| Confident | 20,168 | 29.1 | 1,124 | 31.7 | .001 | (1) |
| Not confident | 49,169 | 70.9 | 2,425 | 68.3 |  |  |
| **Concerns about Abusing Own Child** |  |  |  |  |  |  |
| No | 52,431 | 75.6 | 2,597 | 74.5 | .126 | (1) |
| Yes | 16,906 | 24.4 | 890 | 25.5 |  |  |

Note. Differences in all variables between the included and excluded ones were estimated using the $\chi^2$ tests
\**p* < .05

**Appendix 2.** Results of Logistic Regression Analysis with “Isolation of Child-Rearing” as the Objective Variable (MICE)

|  | OR | Multivariate analysis<br>95% CI | *P* * |
| --- | --- | --- | --- |
| **Maternal Age at Childbirth** |  |  |  |
| ≤ 24 years | 1.2 | [0.7, 2.0] | .554 |
| 25–29 years | 1.3 | [0.9, 1.9] | .210 |
| 30–34 years | ref |  |  |
| 35–39 years | 1.6 | [1.0, 2.4] | .029 |
| ≥ 40 years | 2.2 | [1.1, 4.4] | .019 |
| **Child Age** |  |  |  |
| 3–4 months | ref |  |  |
| 18 months | 1.0 | [0.6, 1.6] | .999 |
| 3 years | 1.6 | [1.0, 2.5] | .033 |
| **Mothers' Employment Status** |  |  |  |
| Employed | ref |  |  |
| Unemployed | 1.7 | [1.2, 2.3] | .001 |
| **Socio-Economic Status** |  |  |  |
| Average or above average | ref |  |  |
| Difficult | 1.9 | [1.4, 2.5] | < .001 |
| **Husband's Participation in Child-Rearing** |  |  |  |
| Yes | ref |  |  |
| Almost none | 5.6 | [4.1, 7.8] | < .001 |
| **Living Municipality** |  |  |  |
| City | ref |  |  |
| City designated by | 1.2 | [0.7, 2.2] | .552 |
| Town/village | 1.0 | [0.6, 1.5] | .874 |
| Special ward (Tokyo ward) | 3.9 | [2.0, 7.5] | < .001 |
| **Time to Relax with Child** |  |  |  |
| Yes | ref |  |  |
| No | 4.8 | [3.3, 7.0] | < .001 |
| **Child-Rearing Confidence** |  |  |  |
| Confident | ref |  |  |
| Not confident | 1.4 | [0.9, 2.3] | .169 |
| **Concerns about Abusing Own Child** |  |  |  |
| No | ref |  |  |
| Yes | 2.0 | [1.4, 2.7] | < .001 |

\**p* < .05

**Appendix 3.** Frequencies of Complete and Imputed Variables.

|  | Complete | Imputed | Total |
| --- | --- | --- | --- |
| **Maternal Age at Childbirth** | 75,277 | 345 | 75,622 |
| **Child Age** | 75,622 | 0 | 75,622 |
| **Mothers' Employment Status** | 73,070 | 2,552 | 75,622 |
| **Socio-Economic Status** | 73,037 | 2,585 | 75,622 |
| **Husband's Participation in Child-Rearing** | 71,019 | 4,603 | 75,622 |
| **Living Municipality** | 75,622 | 0 | 75,622 |
| **Time to Relax with Child** | 73,018 | 2,604 | 75,622 |
| **Child-Rearing Confidence** | 72,886 | 2,736 | 75,622 |
| **Concerns about Abusing Own Child** | 72,824 | 2,798 | 75,622 |

## References

Brashears, M. E. (2011). Small networks and high isolation? A re-examination of American discussion networks. Social Networks, 33 (4), 331–341. https://doi.org/10.1016/j.socnet.2011.10.003

Cabinet Office. (2004). 2004 Edition Declining Birthrate White Paper 2004 (full version). https://www8.cao.go.jp/shoushi/shoushika/whitepaper/measures/w-2004/html_h/index.html

Cabinet Office. (2018). Initiatives of promotion project of employees using paternity leave (Ikumen Project). Gender Equality, 6, 2–5. http://www.gender.go.jp/public/kyodosankaku/2018/201806/pdf/201806.pdf

Chou, K. L., Liang, K., & Sareen, J. (2011). The association between social isolation and DSM-IV Mood, Anxiety, and Substance use disorders: Wave 2 of the national epidemiologic survey on alcohol and related conditions. The Journal of Clinical Psychiatry, 72 (11), 1468–1476. https://doi.org/10.4088/JCP.10m06019gry

Eckert, S., & Kohler, S. (2014). Urbanization and health in developing countries: A systematic review. World Health & Population, 15 (1), 7–20. https://doi.org/10.12927/whp.2014.23722

Elovainio, M., Hakulinen, C., Pulkki-Råback, L., Virtanen, N., Josefsson, K., Jokela, M., Kivimäki, M. (2017). Contribution of risk factors to excess mortality in isolated and lonely individuals: An analysis of data from the UK Biobank cohort study. Lancet Public Health, 2 (6), e260–e266. https://doi.org/10.1016/S2468-2667(17)30075-0

Gallie, D., Paugam, S., & Jacobs, S. (2003). Unemployment, poverty and social isolation: Is there a vicious circle of social exclusion? European Societies, 5 (1), 1–32. https://doi.org/10.1080/1461669032000057668

Gordon, D., Levitas, R., Pantazis, C., Patsios, D., Payne, S., Toensent, P., & Williams, J. (2006). >Poverty and Social Exclusion in Britain. Joseph Rowntree Foundation.

Holt-Lunstad, J., Smith, T. B., Baker, M., Harris, T., & Stephenson, D. (2015). Loneliness and social isolation as risk factors for mortality: A meta-analytic review. Perspectives on Psychological Science, 10 (2), 227–237. https://doi.org/10.1177/1745691614568352

Honda, Y., Fujiwara, T., & Kawachi, I. (2019). Higher child-raising costs due to maternal social> isolation: Large population-based study in Japan. Social Science and Medicine Journal, 233, 71–77. https://doi.org/10.1016/j.socscimed.2019.06.001

International Review of Leave Policies and Related Research. (2019). International Network on Leave Policies and Research. https://www.leavenetwork.org/fileadmin/user_upload/k_leavenetwork/annual_reviews/2019/2._2019_Compiled_Report_2019_0824-.pdf

Kennedy, D. P., & Adolphs, R. (2011). Social neuroscience: Stress and the city. Nature, 474 (7352), 452–453. https://doi.org/10.1038/474452a

Kim, T. H., Rotondi, M., Connolly, J., & Tamim, H. (2017). Characteristics of social support among teenage, optimal age, and advanced age women in Canada: An analysis of the national longitudinal survey of children and youth. Maternal and Child Health Journal, 21 (6), 1417–1427. https://doi.org/10.1007/s10995-016-2249-9

Krabbendam, L., & Van, O. (2005). Schizophrenia and urbanicity: A major environmental influence—conditional on genetic risk. Schizophrenia Bulletin, 31 (4), 795–799. https://doi.org/10.1093/schbul/sbi060

Lederbogen, F., Kirsch, P., Haddad, L., Streit, F., Tost, H., Schuch, P., & Meyer-Lindenberg, A. (2011). City living and urban upbringing affect neural social stress processing in humans. Nature, 474 (7352), 498–501. doi: 10.1038/nature10190

Leigh-Hunt, N., Bagguley, D., Bash, K., Turner, V., Turnbull, S., Valtorta, N., & Caan, W. (2017). An overview of systematic reviews on the public health consequences of social isolation and loneliness. Public Health, 152, 157–171. https://doi.org/10.1016/j.puhe.2017.07.035

Lindsay, C. (2010). In a lonely place? Social networks, job seeking and the experience of long-term unemployment. Social Policy and Society, 9 (1), 25–37. https://doi.org/10.1017/S1474746409990170

Maternal and Child Health Division, Children and Families Bureau, Ministry of Health, Labour and Welfare. (2000). Meeting Report, Healthy Parents and Children 21. https://www.mhlw.go.jp/www1/topics/sukoyaka/tp1117-1_c_18.html

Ministry of Health, Labour and Welfare. (2005, March 25). A Guide How to Handle Child Abuse. https://www.mhlw.go.jp/bunya/kodomo/dv05/00.html

Mitsubishi UFJ Research and Consulting. (2015). 2015 Research Project Report on the State of Support for Work-Life Balance: Worker Survey Results. https://www.mhlw.go.jp/file/06-Seisakujouhou-11900000-Koyoukintoujidoukateikyoku/0000103116.pdf

Mochizuki, Y., Tanaka, E., Shinohara, R., Sugisawa, Y., Tomisaki, E., Watanabe, T., Anme, T. (2014). The influence of caregivers’ anxiety and the home environment on child abuse: A study of children attending child-care centers. Japanese Journal of Public Health, 61 (6), 263–274. https://doi.org/10.11236/jph.61.6_263

Mulvaney, C., & Kendrick, D. (2005a). Depressive symptoms in mothers of pre-school children. Effects of deprivation, social support, stress and neighbourhood social capital. Social Psychiatry and Psychiatric Epidemiology, 40 (3), 202–208. https://doi.org/10.1007/s00127-005-0859-4

Mulvaney, C., & Kendrick, D. (2005b). Do maternal depressive symptoms, stress and a lack of social support influence whether mothers living in deprived circumstances adopt safety practices for the prevention of childhood injury? Child Care, Health and Development, 32 (3), 311–2319. https://doi.org/10.1111/j.1365-2214.2006.00590.x

OECD Family Database. (2019). Age of mothers at childbirth and age-specific fertility 2019. https://www.oecd.org/els/soc/SF_2_3_Age_mothers_childbirth.pdf

Osawa, E., Ojima, T., Akiyama, Y., & Ymagata, Z. (2019). National campaign maternal and child health in 21^st^-century Japan: Healthy Parents and Children 21. Journal of the National Institute of Public Health, 68 (1), 2–7. https://doi.org/10.20683/jniph.68.1_2

Pedersen, C. B., & Mortensen, P. B. (2001). Evidence of a dose-response relationship between urbanicity during upbringing and schizophrenia risk. Archives of General Psychiatry, 58 (11), 1039–1046. https://doi.org/10.1001/archpsyc.58.11.1039

Sarnoff, I., & Zimbardo, P. G. (1961). Anxiety, fear, and social isolation. The Journal of Abnormal and Social Psychology, 62 (2), 356–363. https://doi.org/10.1037/h0046506

Stewart, M. J., Makwarimba, E., Reutter, L. I., Veenstra, G., Raphael, D., & Love, R. (2009). Poverty, sense of belonging and experiences of social isolation. Journal of Poverty, 13 (2), 173–195. https://doi.org/10.1080/10875540902841762

Strange, C., Fisher, C., Howat, P., & Wood, L. (2014). The essence of being connected: The lived experience of mothers with young children in newer residential areas. Community Work & Family, 17 (4), 486–502. https://doi.org/10.1080/13668803.2014.935704

U.S. Department of Health and Human Services. (2019). 2019 Prevention Resource Guide. Retrieved from: https://www.childwelfare.gov/pubPDFs/guide_2019.pdf

UNICEF Office of Research - Innocenti. (2019a). Are the world’s richest countries family friendly? Policy in the OECD and EU. https://www.unicef-irc.org/publications/pdf/Family-Friendly-Policies-Research_UNICEF_%202019.pdf. pp.10

UNICEF. (2019b). Paid Parental Leave and Family-Friendly Policies: An evidence brief. https://www.unicef.org/sites/default/files/2019-07/UNICEF-Parental-Leave-Family-Friendly-Policies-2019.pdf

White, I. R., Royston, P., & Wood, A. M. (2011). Multiple imputation using chained equations: Issues and guidance for practice. Statistics in Medicine, 30 (4), 377–399. http://doi.org/10.1002/sim.4067

Yamagata, Z., Matsuura, K., Yamazaki, Y., Ojima, T., Tamakoshi, K., Ichikawa, K., & Akiyama, Y. (2014). Study on final survey and analyzing problems of Healthy Parents and Children 21, and promotion of the next national health movement: Summary and division reports. http://sukoyaka21.jp/expert/yamagata-report

Yamazaki, S., Shinohara, R., Akiyama, Y., Ichikawa, K., Ojima, T., Tamakoshi, K., & Yamagata, Z. (2018). The relationship between parenting anxiety in mothers and the resources from which they routinely sought advice: The final “Healthy Parents and Children 21” survey. Japanese Journal of Public Health, 65 (7), 334–346. https://doi.org/10.11236/jph.65.7_334

